# MechaScreener: Large Language Model-Based Automated Screening for Systematic Reviews and Research

**DOI:** 10.64898/2026.04.28.26352009

**Authors:** Connor Forbes, Matt Carter, Carly Hudson, Paul Glasziou, Justin Clark

## Abstract

Systematic Reviews (SRs) are the gold standard for evidence synthesis, but the manual title and abstract screening of thousands of references creates a severe bottleneck. Existing automated tools have historically struggled to achieve the near-perfect recall (sensitivity) required for reliable reviews. We developed MechaScreener as a “zero-shot” automated screening tool that utilises a Large Language Model (LLM) to rank article relevance. The tool requires no initial training data or manual pre-screening, as MechaScreener directly applies user-provided question elements (PICO) or inclusion/exclusion criteria to assign an inclusion probability score (1−5) to each reference. We evaluated the tool in two phases: a development phase using five reference libraries to optimise prompts, and an independent evaluation phase using 10 diverse Cochrane review libraries (comprising both randomised controlled trials and non-RCTs) containing over 58,000 references. In the evaluation dataset, MechaScreener achieved a perfect mean recall of 1.00 (100%, pooled 95% CI: 0.98-1.00), ensuring no relevant articles were missed. Concurrently, it achieved an overall mean specificity of 0.61 (61%, pooled 95% CI: 0.59-0.60). Specificity varied: from 0.21 in broad public health topics to 0.91 in precise pharmacological interventions-reflecting the tool’s built-in conservatism when evaluating ambiguous abstracts. By safely eliminating over 60% of irrelevant literature during the initial screening phase without compromising recall, MechaScreener functions as a highly reliable but low-effort “first-pass” filter, allowing researchers to substantially reduce manual workloads and reallocate resources toward full-text review and data extraction.

## 1 Introduction

Systematic Reviews (SRs) are considered the ideal method to summarise current research on a specific research question; however, they are time-consuming, resource-heavy and can be out of date by the time they are published. Systematic reviews require multiple, complex tasks to produce [1]. One of the first tasks in an SR, screening search results, often requires the review of thousands of irrelevant references [2], which must be manually screened by at least two reviewers [3]. This process is time-consuming: Bramer et al. [4] estimated a rate of 60-300 references per hour. Variation in screening speed can be attributed to team experience, the inclusion and exclusion criteria, and the software used.

Automation tools might reduce this burden by assessing the relevance of references for a given review, providing researchers with a metric to quickly ascertain whether a study is likely or unlikely to meet inclusion criteria. Most current tools have used machine learning - which requires a training set; hence, the reviewers must manually screen a subset of several hundred references to train the algorithm (e.g., active learning models such as Abstrackr, ASReview, and Rayyan [5]). More recently, Large Language Models (LLMs) (e.g., OpenAI’s GPT models, Anthropic’s Claude, Alibaba’s Qwen series) have emerged as a promising alternative to achieve this; however, recall continues to be a limiting factor [6]. Recent evaluations of LLMs in literature screening indicate that while a recall of 1.00 (100%) is possible, it usually requires specific settings utilising ensemble models and probability calibration, albeit frequently at the cost of reduced precision [7]. To effectively automate screening, recall (the number of relevant studies included) needs to be near or at 1.00 (meaning all relevant studies were correctly identified). To date no published LLM screening method has been able to achieve a recall of 1.00 consistently across reviews, while reducing the screening burden in a significant way [8].

To reduce the screening burden, we developed MechaScreener. MechaScreener is an LLM screening tool available via the Evidence Review Accelerator (TERA) [9]. Unlike the current machine learning tools that rely on a manually screened subset of references to train the algorithm, MechaScreener operates as a “zero-shot” tool. This means it does not require any training examples or manual pre-screening. Instead, it relies entirely on the LLM’s pre-existing language capabilities to evaluate references; users simply upload their references and enter their review’s eligibility criteria in plain text. This study outlines the development of MechaScreener and its screening results across 15 systematic reviews.

## 2 Methods

### 2.1 System Architecture

MechaScreener development started in 2024 using a Node.js/TypeScript backend environment with a Vue 3 frontend. The system processes reference citations in batches, extracting the title and abstract and pairing them with a user-provided screening criteria string; formatted as Population, Intervention, Comparator, Outcome, Type of study (PICOT) criteria [10] or general inclusion/exclusion rules.

### 2.2 Prompting and Scoring

The MechaScreener system utilises a structured prompt consisting of:

1. **System Prompt:** Defined instructions for the **LLM** to act as a screening assistant.
2. **Reference Data:** Title and abstract of the paper.
3. **Criteria:** The eligibility criteria for the specific review.

In the context of Large Language Models, `temperature` is a parameter that controls the randomness or creativity of the model’s output distribution, while `top_k` restricts the model to sample only from the *k* most probable next tokens. To ensure the screening process is completely deterministic and highly reproducible for research purposes, we utilised a model configuration with a `temperature` of O and `top_k` of 1, effectively forcing the model to always select the single most likely response.

The model is instructed to provide a score from 1 (low probability of inclusion) to 5 (high probability of inclusion).

### 2.3 Experimental Design

The evaluation of MechaScreener was conducted in two phases:

- **Development Dataset:** Five control reference libraries were used to calibrate the prompt and scoring threshold aimed at maximising recall over specificity.
- **Evaluation Dataset:** Ten additional reference libraries were used for independent validation of the tool’s performance across different medical and social science domains (e.g., COVID-19, diabetes, bone health). We included five SRs focused on Randomised Controlled Trials (RCTs) and five SRs focused on non-RCT studies. All 15 reference libraries are available online in our GitHub repository.

The eligibility criteria prompts provided to the LLM were developed by three individuals with varying levels of systematic review experience (a novice reviewer, a PhD student, and an experienced reviewer), with each individual designing the prompts for five libraries. The exact eligibility criteria provided to the LLM for each library are detailed in Supplementary File 1. This file can also be used as a prompt guide for using MechaScreener. The 10 evaluation studies were sourced from previous Cochrane reviews. As title/abstract screening results are not available, we used the final full-text inclusion decisions to determine what the relevant studies were.

To compile the set of reviews for the evaluation, we reran searches from 10 randomly selected Cochrane reviews. The sampling frame was created by searching PubMed for a Cochrane review, then 10 were selected by generating a random number (using the Google random number generator) between one and the total number of search results found. For example, if 1000 results were found, the random number was between 1 and 1000. The search result that corresponded to this number was checked to see if it met our inclusion criteria of 1) having searched three or more databases; 2) reporting search strategies for all databases; 3) having an identifiable list of included studies; 4) having a minimum of 6 to a maximum of 50 included studies; and 5) for five of the reviews, including randomised controlled trials, and for the other five, including study types other than randomised controlled trials. Any references missing abstract data were excluded from the evaluation.

### 2.4 Metrics

The performance of MechaScreener is evaluated primarily using two metrics: recall (sensitivity) and specificity (selectivity).

#### Recall (or sensitivity)

measures the proportion of actual relevant studies that were correctly identified by the tool. In the context of systematic reviews, maximising recall is paramount to ensure no critical literature is missed. It is defined as:

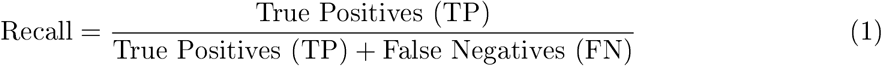

#### Specificity (or selectivity)

measures the proportion of irrelevant studies that were correctly excluded. A higher specificity indicates a greater reduction in the manual screening workload, as fewer irrelevant articles are passed to the human reviewers. It is defined as:

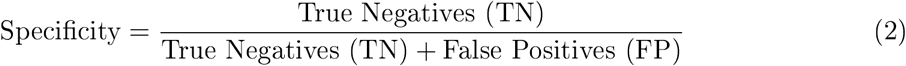

#### Wilson Score Confidence Interval (95% CI)

is used to calculate the confidence bounds for recall and specificity. Standard Wald confidence intervals fail mathematically when proportions reach 0% or 100% (as seen with perfect recall). The Wilson Score Interval is robust for these extreme values [11]. It is calculated as:

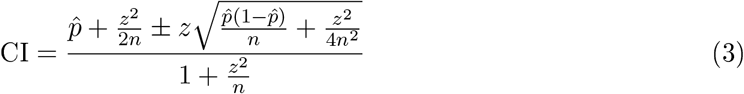

where 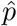 is the observed proportion, *n* is the total number of observations, and *z* is the z-score (1.96 for a 95% confidence level).

### 2.5 Development Results

Five control libraries were used for development to maximise recall (number of references correctly included) and specificity (number of references correctly excluded). These libraries were compiled based on previous reviews conducted at the Institute for Evidence-Based Healthcare (IEBH). The developmental results for these libraries are presented in Table 1.

**Table 1:** Results Across Five Development Libraries.

| Library Name | No. of Refs | Include Studies Correctly Included | Exclude Studies Correctly Excluded |
| --- | --- | --- | --- |
| Antibiotic Prescribing and Tele-health [12] | 934 | 13/13 (100%) | 612/921 (66%) |
| Long Covid [13] | 1,216 | 16/16 (100%) | 552/1,200 (46%) |
| Natural History Primary Care [14] | 5,913 | 11/11 (100%) | 3,169/5,902 (54%) |
| Non Drug Interventions [15] | 5,342 | 35/35 (100%) | 3,972/5,307 (75%) |
| Salt Substitution [16] | 889 | 23/23 (100%) | 733/866 (85%) |

To maximise the recall score, the threshold was optimised to the highest possible score that still maintained a perfect recall of 1.0. An LLM was used to rank each article on a scale from one to five. Empirically, setting the inclusion threshold in the development set at a score of two or higher and excluding only the articles that scored a one, achieved the target 1.00 recall while preserving a reasonable degree of specificity. This prioritised finding all relevant evidence so that the researcher can focus on making fine-grained decisions about what to exclude.

### 2.6 Evaluation Dataset

The evaluation set for the performance of MechaScreener used 10 reference libraries that included a diverse range of topics shown below (five studies that were RCTs and five studies that were non-RCTs).

## 3 Results

MechaScreener was tested across 10 evaluation libraries containing a total of over 58,000 references. Figure **1** summarises the performance metrics visually across each library and highlights the overall average for the evaluation set.

**Figure 1:**
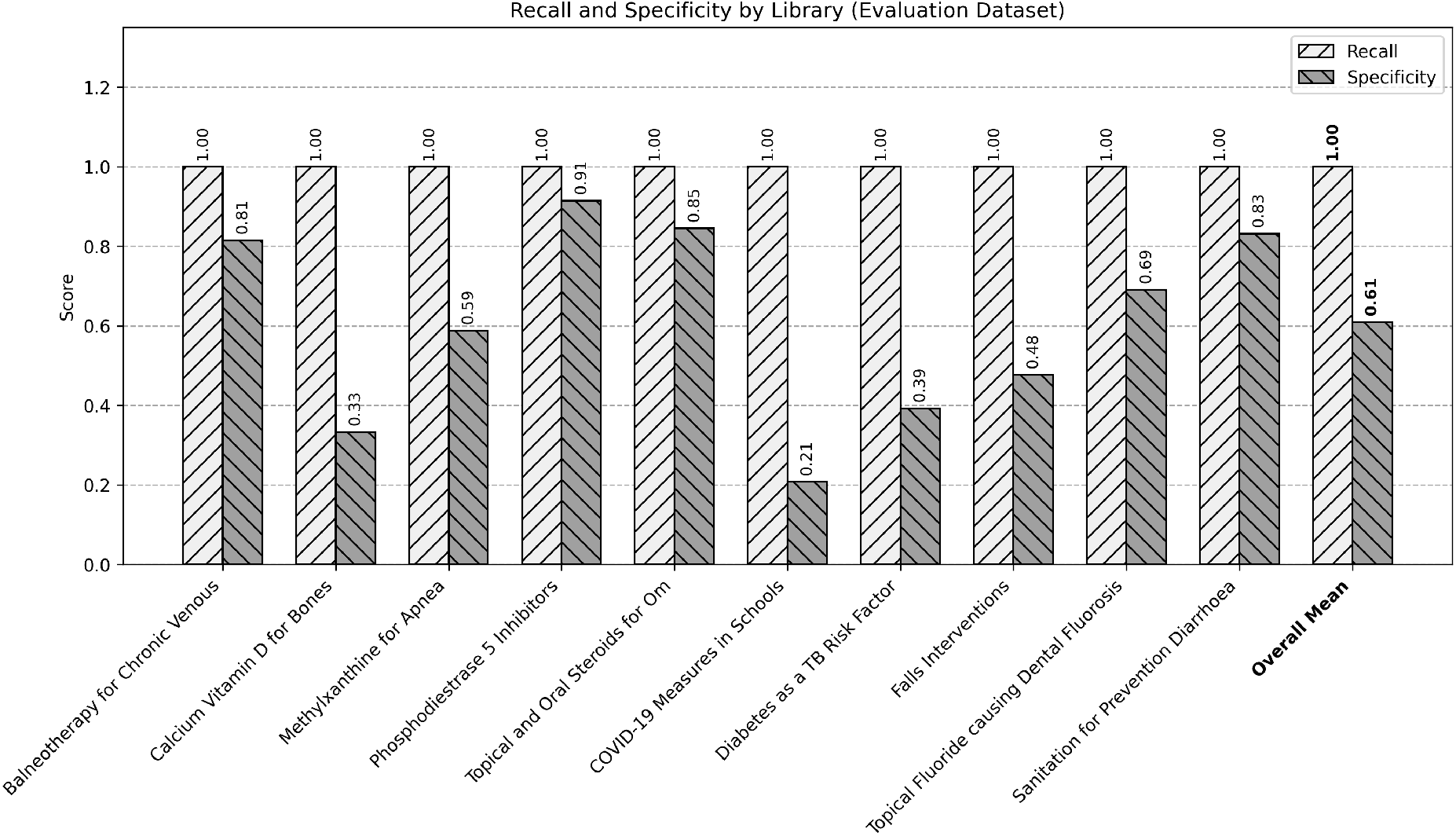
Recall and Specificity Across 10 Evaluation Libraries, Including the Overall Mean.

Within the evaluation dataset, the system achieved a perfect mean recall score (1.00, pooled 95% CI: 0.98-1.00), meaning no relevant articles were missed by the tool. The mean specificity was 0.61 (pooled micro-average 0.59, 95% CI: 0.59-0.60), indicating that MechaScreener correctly excluded 61% of irrelevant articles, significantly narrowing the pool for human review.

An individual breakdown of each evaluation library is presented in Table 3.

We calculated the 95% Wilson Score Confidence Intervals. Table 4 presents these statistical measurements, alongside the macro-average (mean of proportions) and micro-average (pooled data) across all evaluation datasets.

## 4 Discussion

Our results demonstrate that MechaScreener, when prompted with a structured 1-5 scoring system, can act as a highly effective and safe “first-pass” filter across a diverse set of reviews. MechaScreener was able to consistently maintain a perfect mean recall (1.00, pooled 95% CI: 0.98-1.00) across all evaluated libraries while safely removing an average of approximately 61% of the irrelevant literature (mean specificity of 0.61, pooled 95% CI: 0.59-0.60). This translates to a substantial reduction in the manual screening workload for researchers, reallocating valuable time and human resources towards downstream tasks such as full-text review, data extraction, and risk of bias assessment.

### 4.1 Performance Variance and Specificity

While the recall remained stable at 100% across both the development and evaluation datasets, we observed notable variance in specificity depending on the systematic review topic and study design. This variance demonstrates that the LLM’s capacity to confidently discard irrelevant literature is highly dependent on the precision of the research question and the standardisation of the field’s terminology.

For instance, highly specific pharmacological interventions such as *Phosphodiesterase 5 Inhibitors* [l20] achieved a specificity of 0.91, effectively eliminating over 90% of irrelevant studies. Clinical trials in these domains typically feature highly structured abstracts and unambiguous PICOT elements allowing the LLM to easily detect off-target papers.

Conversely, broader or more complex topics, particularly those involving non-randomised controlled trials, (non-RCTs) like *COVID-19 Measures in Schools* [22] or *Diabetes as a TB Risk Factor* [23], exhibited lower specificities (0.21 and 0.39, respectively). These public health and observational fields, often involve diverse study designs varied outcome measures, and less structured reporting. When abstract information is vague, lacks explicit PICOT details, or touches on related thematic concepts without matching precise criteria, MechaScreener conservatively assigns a higher probability of inclusion. This built-in conservatism prevents the accidental exclusion of relevant studies, correctly prioritising a perfect recall over maximal workload reduction when faced with ambiguity.

### 4.2 Limitations

Despite the robust performance, several limitations must be acknowledged. First, as noted in the methodology, the evaluation dataset’s ground truth for ‘relevant’ studies was based on final full-text inclusion decisions rather than initial human title and abstract screening decisions. Because abstracts often lack complete methodological details, an automated title and abstract screener may correctly flag a study as potentially relevant based on the abstract alone, only for it to be counted as a false positive because the study was ultimately excluded during the human full-text review phase. Consequently, the reported specificity may artificially underestimate the tool’s true precision at the abstract screening level.

Second, the current methodology relies heavily on the availability of study abstracts. As highlighted in Table 2, thousands of references across the tested libraries (e.g., over 2,500 in the *Topical Fluoride causing Dental Fluorosis* library) lacked abstract data. Studies without abstracts are inherently difficult to screen automatically and often default to human review, reducing the overall workload savings.

**Table 2:** Overview of References and Missing Abstracts in Evaluation Libraries.

| Library Name | Total References | Missing Abstracts |
| --- | --- | --- |
| **RCT Studies** |  |  |
| Balneotherapy for Chronic Venous [17] | 338 | 11 |
| Calcium Vitamin D for Bones [18] | 5,756 | 255 |
| Methylxanthine for Apnea [19] | 2,781 | 334 |
| Phosphodiesterase 5 Inhibitors [20] | 2,672 | 338 |
| Topical and Oral Steroids for Om [21] | 4,292 | 583 |
| **Non-RCT Studies** |  |  |
| COVID-19 Measures in Schools [22] | 2,162 | 21 |
| Diabetes as a TB Risk Factor [23] | 5,177 | 165 |
| Falls Interventions [24] | 18,098 | 1,095 |
| Topical Fluoride causing Dental Fluorosis [25] | 11,829 | 2,527 |
| Sanitation for Prevention Diarrhoea [26] | 10,770 | 297 |

**Table 3:** Results Across 10 Evaluation Libraries.

| Library Name | No. of Refs | Include Studies Correctly Included | Exclude Studies Correctly Excluded |
| --- | --- | --- | --- |
| Balneotherapy for Chronic Venous | 327 | 9/9 (100%) | 259/318 (81%) |
| Calcium Vitamin D for Bones | 5,493 | 6/6 (100%) | 1,825/5,487 (33%) |
| Methylxanthine for Apnea | 2,446 | 28/28 (100%) | 1,422/2,418 (59%) |
| Phosphodiesterase 5 Inhibitors | 2,333 | 6/6 (100%) | 2,128/2,327 (91%) |
| Topical and Oral Steroids for Om | 3,706 | 28/28 (100%) | 3,110/3,678 (85%) |
| COVID-19 Measures in Schools | 2,141 | 30/30 (100%) | 439/2,111 (21%) |
| Diabetes as a TB Risk Factor | 5,011 | 48/48 (100%) | 1,948/4,963 (39%) |
| Falls Interventions | 17,000 | 9/9 (100%) | 8,102/16,991 (48%) |
| Topical Fluoride causing Dental Fluorosis | 9,294 | 40/40 (100%) | 6,398/9,254 (69%) |
| Sanitation for Prevention Diarrhoea | 10,473 | 31/31 (100%) | 8,684/10,442 (83%) |

**Table 4:** Statistical Summary Across 10 Evaluation Libraries.

| Library Name | Recall (95% CI) | Specificity (95% CI) |
| --- | --- | --- |
| Balneotherapy for Chronic Venous | 1.00 (0.70-1.00) | 0.81 (0.77-0.85) |
| Calcium Vitamin D for Bones | 1.00 (0.61-1.00) | 0.33 (0.32-0.35) |
| Methylxanthine for Apnea | 1.00 (0.88-1.00) | 0.59 (0.57-0.61) |
| Phosphodiesterase 5 Inhibitors | 1.00 (0.61-1.00) | 0.91 (0.90-0.93) |
| Topical and Oral Steroids for Om | 1.00 (0.88-1.00) | 0.85 (0.83-0.86) |
| COVID-19 Measures in Schools | 1.00 (0.89-1.00) | 0.21 (0.19-0.23) |
| Diabetes as a TB Risk Factor | 1.00 (0.93-1.00) | 0.39 (0.38-0.41) |
| Falls Interventions | 1.00 (0.70-1.00) | 0.48 (0.47-0.48) |
| Topical Fluoride causing Dental Fluorosis | 1.00 (0.91-1.00) | 0.69 (0.68-0.70) |
| Sanitation for Prevention Diarrhoea | 1.00 (0.89-1.00) | 0.83 (0.82-0.84) |
| **Macro-Average (Mean)** | **1.00** | **0.61** |
| **Pooled (Micro-Average)** | **1.00 (0.98-1.00)** | **0.59 (0.59-0.60)** |

Third, MechaScreener does not replace human screening. It functions strictly as a “first-pass” filter, meaning that any results the tool flags as potentially relevant still need to be manually screened by a human reviewer to confirm eligibility. Furthermore, MechaScreener is designed explicitly as a title and abstract screening assistant and does not perform full-text screening, meaning human reviewers must also assess the remaining literature in full detail. Finally, while we utilised a deterministic approach (i.e., `temperature = 0`) to ensure reproducibility, the system’s performance remains fundamentally tied to the quality, clarity, and comprehensiveness of the user-provided eligibility criteria. Poorly defined criteria may lead to lower specificity as the model struggles to differentiate borderline studies.

### 4.3 Future Directions

Future research should explore the integration of MechaScreener directly into standard systematic review workfiows and reference management platforms. Evaluating its performance across a broader range of foundational LLMs, as well as refining the prompt architecture to better handle title-only references, are key next steps. Furthermore, expanding the system’s capabilities to assist with automated full-text screening and data extraction could further streamline the evidence synthesis pipeline. Overall, these findings solidify the role of LLMs as powerful, dependable augmentations to traditional systematic review methodologies.

## 5 Conclusion

MechaScreener provides a reliable method for screening research articles. By leveraging large-scale LLMs and a probabilistic scoring approach, researchers can use MechaScreener to reduce their screening workload by more than half while maintaining the rigorous standards required for systematic literature synthesis.

## Declarations

### Data availability statement

The dataset (relevance scores generated by MechaScreener and ground truth inclusion/exclusion decisions), source code for the analysis, along with the scripts used to generate the figures and tables presented in this paper, are publicly archived and available in the GitHub repository: https://github.com/IEBH/mechascreener-paper.

### Ethics approval statement

Not applicable. This study involves the methodological analysis of previously published literature and software performance, and does not involve human or animal subjects.

