## Supplementary File 1 for "MechaScreener: Large Language Model-Based Automated Screening for Systematic Reviews and Research"

### Supplementary File 1: Search Criteria for Evaluated Libraries

MechaScreener: Large Language Model-Based Automated Screening for Systematic Reviews and Research

#### Introduction

This supplementary file details the exact screening criteria provided to the MechaScreener Large Language Model (LLM) for each of the 15 reference libraries analysed in the main text. The libraries are divided into 5 development libraries and 10 evaluation libraries (5 focusing on Randomized Controlled Trials [RCTs] and 5 focusing on non-RCTs). Criteria were structured either using the Population, Intervention, Comparator, Outcome, and Type of study (PICOT) format, or provided as general free-text inclusion/exclusion rules. Only the "include" criteria are listed below; fields left blank in the system are omitted.

### 1 Development Libraries

#### 1.1 Antibiotic Prescribing and Telehealth

**Format:** PICOT

**Population:**
adult or paediatric patients with a history of a community acquired acute infection (respiratory, urinary, or skin and soft tissue)

**Intervention:**
any type of telehealth consultations (phone or video based)

**Comparator:**
face to face consultations

**Outcome:**
the number of antibiotic prescriptions in each type of consultation, follow-up visit rates, testing rates or number of samples sent to the laboratory, any reported adverse events

**Type of study:**
randomized trials or observational studies

#### 1.2 Long COVID

**Format:** PICOT

**Population:**
people of all ages who were eligible to receive a covid-19 vaccine

**Intervention:**
any dose of a covid-19 vaccine

**Comparator:**
no vaccination, an active non-covid-19 vaccine control (eg, influenza vaccine), or placebo

**Outcome:**
patients with a diagnosis of long covid, prevalence of individual symptoms of long covid, such as prolonged fatigue, shortness of breath, cognitive difficulties, and loss of sense of smell

#### 1.3 Natural History of Primary Care

**Format:** General/Other
Eligibility criteria were: clinical practice guidelines produced by an international or national organisation involved in the publication of guidelines, and contained clinical recommendations on the management of acute infections that are commonly seen in primary care and may be managed with antibiotics (including acute respiratory infections (ARIs), lower urinary tract infections (UTIs), skin and soft tissue infections (SSTIs), conjunctivitis).

Inclusion criteria:

1. Acute respiratory infections including acute otitis media, sore throat, pharyngitis, tonsillitis, common cold, sinusitis, laryngitis, acute cough, acute bronchitis, bronchiolitis middle ear inflammatory conditions
2. Rhinosinusitis, rhinitis, group A streptococcal infections, streptococcal pharyngitis
3. Influenza-like illnesses including seasonal influenza

4. Lower urinary tract infections, cystitis, acute bladder infection, recurrent UTI
5. Skin and soft tissue infections, cellulitis, acne, impetigo, boils, carbuncles, ecthyma
6. All forms of conjunctival conditions
7. Management of croup
8. Urinary tract infections in non-pregnant women
9. Acute exacerbation of COPD

##### 1.4 Non-Drug Interventions

**Format:** General/Other
Include a study if it meets ALL of the following criteria:

1. **Study Type:** The study must be a systematic review or a scoping review.
2. **Topic:** It must identify the barriers or enablers of using non-drug interventions (e.g., diet, exercise, psychological support, self-management).
3. **Condition:** The intervention must be for the prevention or management of a chronic disease.
4. **Setting:** The setting must be primary care, where a clinician referred the patient to the intervention.
5. **Data Source:** The review must be based on primary studies that are mostly qualitative or mixed-methods.

##### 1.5 Salt Substitution

**Format:** PICOT

**Population:**
adults (aged $\geq$18 years)

**Intervention:**
studies of salt substitution interventions (that is, salt substitute intake compared with regular salt intake) delivered through advice and recommendation from a clinician or researcher, or the provision of salt substitute product

**Comparator:**
studies that used no intervention or regular table salt as a comparator

**Outcome:**
mortality, MACE (for example, any composite of myocardial infarction, revascularization, stroke, and heart failure), serious adverse events (for example, renal failure or severe hyperkalemia), SBP and DBP

**Type of study:**
randomized trials

#### 2 Evaluation Libraries (RCTs)

##### 2.1 Balneotherapy for Chronic Venous Insufficiency

**Format:** PICOT

**Population:**
people with venous insufficiency

**Intervention:**
patients treated with balneotherapy

**Outcome:**
Edema OR Leg Ulcer OR Pain OR Skin Pigmentation OR Adverse Events OR Quality of Life

**Type of study:**
randomized trials

##### 2.2 Calcium and Vitamin D for Increasing Bone Mineral Density in Premenopausal Women

**Format:** PICOT

**Population:**
Adult women

**Intervention:**
People given Calcium or Vitamin D

**Outcome:**
Increased bone mineral density or reduction in fractures or quality of life improvements or adverse events

**Type of study:**
randomized trials

##### 2.3 Methylxanthine for the Prevention and Treatment of Apnea in Preterm Infants

**Format:** PICOT

**Population:**
Preterm infants

**Intervention:**
People given Methylxanthine or Aminophylline or Theophylline or Caffeine

**Outcome:**
Positive Pressure Therapy or Chronic Lung Disease or Apnea or Death or Neurodevelopmental Disorder

**Type of study:**
randomized trials

##### 2.4 Phosphodiesterase 5 Inhibitors (PDE5i) for the Treatment of Raynaud's Phenomenon

**Format:** PICOT

**Population:**
Adults with Raynaud's Phenomenon

**Intervention:**
People given Phosphodiesterase 5 Inhibitors

**Outcome:**
Frequency of attacks or Duration of attacks or Severity of attacks or Raynaud's condition score or Pain or Adverse Events OR Patient Improvement

**Type of study:**
randomized trials

##### 2.5 Topical and Oral Steroids for Otitis Media with Effusion (OME) in Children

**Format:** PICOT

**Population:**
Children with Otitis Media with effusion

**Intervention:**
People given steroid therapy

**Outcome:**
Hearing or Quality of Life or Adverse Events or persistence of otitis media

**Type of study:**
randomized trials

#### 3 Evaluation Libraries (Non-RCTs)

##### 3.1 COVID-19 Measures in Schools

**Format:** PICOT

**Population:**
people involved in school settings

**Intervention:**
measures to reduce transmission of COVID-19

**Outcome:**
"transmission-related outcomes" OR "healthcare utilisation" OR "economic outcomes" OR "social outcomes" OR "ecological outcomes"

##### 3.2 Diabetes as a TB Risk Factor

**Format:** PICOT

**Population:**
patients with diabetes

**Outcome:**
incidence of tuberculosis

**Type of study:**
cohort studies

##### 3.3 Falls Interventions

**Format:** PICOT

**Population:**
patients over 45 years old

**Intervention:**
"fall prevention" OR "exercise" OR "safety education" OR "nutrition therapy"

**Comparator:**
usual care

**Outcome:**
"incidence of falls" OR "hospitalisations for falls" OR "fall-related injuries" OR "cost of healthcare associated with falls"

**Type of study:**
"randomised controlled trials" OR "step-wedge designs" OR "non-RCTs"

##### 3.4 Topical Fluoride causing Dental Fluorosis

**Format:** PICOT

**Population:**
children exposed to topical fluoride

**Intervention:**
topical fluoride

**Comparator:**
"non-topical fluoride treatments" OR "placebo" OR "no intervention"

**Outcome:**
prevalence of fluorosis

**Type of study:**
"randomised controlled trials" OR "observational studies" OR "cohort studies" OR "case control studies" OR "cross-sectional surveys"

##### 3.5 Sanitation for Prevention Diarrhoea

**Format:** PICOT

**Population:**
"children" OR "adults"

**Intervention:**
sanitation measures

**Comparator:**
"open defecation" OR "current practice"

**Outcome:**
prevalence of diarrhoea

**Type of study:**
"randomised controlled trials" OR "quasi-RCTs" OR "non-RCTs" OR "controlled before and after studies" OR "matched cohort studies"
